## Supplementary Materials for "Impact of the COVID-19 pandemic on community antibiotic prescribing and stewardship: a qualitative interview study with general practitioners in England"

Some planning related to current priorities and potential changes to infection management ... 23

### Supplementary Material 1: Interview topic guides

Three interview guides were used:

(1) For GPs in the STEP-UP study in autumn 2020 (late October to early December 2020);
(2) For additional GPs in autumn 2020;
(3) For all GPs in spring 2021 (late April to early June 2021).

#### (1) Topic guide for GPs participating in the STEP-UP study (autumn 2020)

*Questions related to the impact of the Covid-19 pandemic:*

1. Could you tell me about how your practice has been affected by the Covid-19 pandemic?
   *Prompts, e.g.:*
   a. How have your practice and consultations changed since Covid-19?
   b. How have things changed since the start of the pandemic?

2. To what extent has the pandemic affected how you view or approach antibiotic prescribing and antimicrobial stewardship?
   *Prompts, e.g.:*
   a. How has the situation influenced how likely you are to prescribe antibiotics?
   b. How do you feel the pandemic might have influenced antibiotic prescribing rate in your practice?
   c. Have you reviewed antibiotic prescribing data for your practice since March/April?

3. Have you or your practice stopped or taken up any initiatives to optimise antibiotic prescribing since the start of the pandemic?

*Questions related to the STEP-UP study (and AMS strategies promoted as part of it):*

4. To what extent has the way you talk about antibiotics been influenced by Covid-19?
5. How has the use of leaflets been influenced by Covid-19?
6. How has point-of-care CRP testing been affected by Covid-19?
7. Given remote consultations may continue in general practice for some time, how do you think point-of-care testing could be incorporated to guide treatment decisions?
8. To what extent has the pandemic influenced whether and how you use delayed/back-up prescriptions?
9. Questions related to a website about AMS strategies provided as part of the STEP-UP study.

*Questions about implications:*

10. What do you think are the implications of the pandemic on antibiotic prescribing and antimicrobial stewardship?
    *Prompts, e.g.:*
    a. What do you think will be the long-lasting effects of the pandemic on general practice and AMS?

11. What do you think would help you and your practice to prescribe antibiotics prudently during the pandemic? What kind of support would be helpful? Who could provide it?

12. In general, do you have any thoughts on, or suggestions for, how we may better support appropriate antibiotic prescribing in the future?

13. Is there anything else that you'd like to share with me?

### (2) Topic guide for additional GPs (autumn 2020)

*Questions related to the impact of the Covid-19 pandemic:*

1. Could you tell me about how your practice has been affected by the Covid-19 pandemic?
   *Prompts, e.g.:*
   a. How have your practice and consultations changed since Covid-19?
   b. How have things changed since the start of the pandemic?
   c. How has the pandemic impacted on general practice professionals?
   d. How do you think has it impacted on patients?

2. How do you think the pandemic might have influenced your antibiotic prescribing? (or how likely you are to prescribe antibiotics?)
   a. Has it influenced how you use immediate or delayed prescriptions?
   b. To what extent has your antibiotic prescribing for patients with RTIs / suspected Covid-19 / other infections been influenced?
   c. How has the pandemic influenced how you view or approach antibiotics as a practice?
   d. Have you reviewed antibiotic prescribing data for your practice since March/April?
   e. Do you know how antibiotic prescribing rates have changed during the pandemic?
   f. How do you feel the pandemic might have influenced antibiotic prescribing rate in your practice?

3. To what extent has the pandemic affected how you view or approach antimicrobial stewardship, and what you did or wanted to do, if anything, as part of it?
   a. Have you or your practice stopped any initiatives to optimise antibiotic prescribing since the start of the pandemic? Have you taken up any initiatives?
   b. To what extent has the way you talk about antibiotics been influenced by Covid-19?
   c. How has the use of leaflets been influenced by Covid-19?
   d. To what extent has the pandemic influenced whether and how you use delayed/back-up prescriptions?
   e. How do you currently issue delayed prescriptions? Has that changed during the pandemic?
   f. Do you have access to point-of-care CRP testing in your practice? (If yes – ask about it, if not – ask about general views on it)
   g. How has the use of point-of-care (CRP) testing been affected by the pandemic?
   h. Given remote consultations may continue in general practice for some time, how do you think point-of-care testing could be incorporated to guide treatment decisions?

4. Since the start of the pandemic, have you used any resources or tools to help you prescribe antibiotics appropriately? (E.g. any training, websites, guidelines, clinical scores)
   a. [If yes] How did you use these resources/tools?
   b. How useful or not have they been to you?
   c. What prevented or stopped you from using such resources/tools?

*Questions about implications:*

5. What do you think are the implications of the pandemic on antibiotic prescribing and antimicrobial stewardship?
   *Prompts, e.g.:*
   a. What do you think would help you (and your practice) to prescribe antibiotics prudently during the pandemic? What kind of support would be helpful and who could provide it?
   b. Do you have any thoughts on, or suggestions for, how we may generally better support appropriate antibiotic prescribing in the future?

6. Is there anything else that you'd like to share with me?

### (3) Topic guide for all GPs (spring 2021)

*Questions related to the impact of the Covid-19 pandemic:*

1. Could you tell me about the main ways how your practice has been affected by the Covid-19 pandemic (since last interview)?
   *Prompts, e.g.:*
   a. How have your practice and consultations changed since autumn?
   b. (*If not interviewed previously*) How have things changed throughout the year since the start of the pandemic?
   c. How is doing the Covid-19 vaccinations impacting on your practice?
   d. How has the pandemic impacted on general practice professionals?
   e. How do you think has the pandemic impacted on your patients?

2. How do you think the pandemic might have influenced your antibiotic prescribing? (Or how likely you are to prescribe antibiotics?) (since autumn, or throughout the year)
   *Prompts, e.g.:*
   a. To what extent has your antibiotic prescribing for patients with RTIs/ Covid-19/ other infections been influenced?
   b. How would you manage these patients if seeing them in-person rather than consulting remotely?
   c. Do you know how antibiotic prescribing rates in your practice have changed during the pandemic? (compared to pre-pandemic) Why do you think that might be?
   d. Have you reviewed antibiotic prescribing data for your practice in 2020?
   e. How do you feel the pandemic might have influenced antibiotic prescribing rate in your practice?

3. To what extent has the pandemic influenced how you view or approach AMS?
   *Prompts, e.g.:*
   a. Have you or your practice stopped any initiatives to optimise antibiotic prescribing since autumn / since the start of the pandemic?
   b. Have you or your practice taken up any initiatives since the start of the pandemic?
   c. To what extent has the pandemic influenced how you view or approach antibiotics as a practice?

4. Strategies: Since autumn/the start of the pandemic, have you used any resources, tools or training related to prescribing or optimising antibiotics?

*Prompts, e.g.:*

a. [If yes] How have you used these? How useful or not have they been to you?
b. [If not] What prevented or stopped you from using such resources/tools?
c. Has your use or engagement with these been different before and since the pandemic?

5. Have you used any strategies to help with appropriate and prudent prescribing of antibiotics?

   *Prompts, e.g.:*

   a. Has the way you talk to patients about antibiotics been influenced by Covid-19? Has it changed since autumn/over the past year? How?
   b. Has the use of patient leaflets been influenced by Covid-19? Has it changed since autumn/over the past year? How?
   c. Has the pandemic influenced how you delayed antibiotic prescriptions? How?
   d. How do you think delayed prescriptions could be incorporated/used in remote consultations going forward?
   e. Do you have access to point-of-care CRP testing in your practice? (If yes – ask about it, if not – ask about general views on it)
   f. How do you think point-of-care (CRP) testing could be incorporated in the future to guide treatment decisions?

*Questions about implications:*

6. What do you think are the future implications of the pandemic on antibiotic prescribing and antimicrobial stewardship? (next autumn/winter, long-term)

7. What are your current priorities?

   *Prompts, e.g.:*

   a. How important is antibiotic optimisation in your practice among the different priorities?
   b. How do you plan to consult in the future?
   c. Who might the remote consultations be used with or not used with in the future? How is it going to affect care?

8. Is there anything that you are planning or would like to do in autumn/2022 to help optimise antibiotic prescribing?
   a. Are you in any discussions with others, e.g. CCG, about planning AMS?
   b. What would help you and your practice to optimise antibiotic prescribing this autumn/in 2022-3?
   c. What kind of support would be helpful and who could provide it?
   d. What would you like to know to help you plan for autumn? What kind of information would help?

9. Do you have any thoughts on, or suggestions for, how we may generally better support appropriate antibiotic prescribing in the future?

10. Is there anything else that you'd like to share with me?

### Supplementary Material 2: Additional quotes supporting the findings

The quotes below were selected from the dataset to further illustrate and support the findings.

#### Fluctuations in antibiotic prescribing during Covid-19

##### *Initial increase in antibiotic prescribing, followed by gradual return to 'normal' prescribing*

Right at the beginning, when COVID first struck, at the beginning of March and everyone went to lockdown and we were still working, it was very much stressed to us we must see patients as an absolute last resort and we should do everything virtually if we could and, therefore, we were treating a lot of chest infections virtually which sounds and goes against everything that we've ever been trained to do but we were told we absolutely mustn't spread COVID, therefore we must keep our distance, our socially, that was when PPE was in very short supply. And talking to my colleagues about it, we certainly were prescribing more antibiotics. We'd never, before COVID, have prescribed antibiotics without seeing a patient and examining them but we were doing that and, I think, talking to other colleagues, that's what everybody was doing. And now as we, well, in the summer, as we approached the restoration phase, we were seeing more patients and examining them so I think our prescribing of antibiotics went down again but now, as we're approaching or now into the second wave, we are very much trying to practise virtually as much as we can as per NHS England advice so, again, potentially prescribing more than we would normally. So it'll be interesting to look at those figures. GP02 Oct '20

I wouldn't be surprised if prescribing has gone up, just because obviously people are trying to cover all eventualities. It's quite, actually if you practice quite a defensive form of medicine, you'd, to be on the safe side you're more likely to give them a course of antibiotics, particularly if you can't really assess them in person. That would be more my gut reaction, I would suggest that it may have slightly increased the prescribing. GP04 Oct '20

I think certainly in the early months of the pandemic antibiotic prescribing was higher than before where we were unsure of the risks of bringing patients into the surgery versus the risks of antimicrobial resistance or the risk of an untreated infection where a lot of people, a lot of prescribers were issuing antibiotics where previously we would have been more conservative and examined the patient first. There were a lot more prescriptions of things like chloramphenicol for conjunctivitis or antibiotics for a chest infection where we would issue them without assessing the patient. Where we were satisfied the patient didn't need admission but unsure whether they needed antibiotics, rather than bring them in to assess, the prescription was simply sent, so I'm sure at the beginning of the pandemic, in sort of April, May, June, lots more antibiotics would have been prescribed than the year before for example. Now I think, now that there are more Covid hubs, I think as a group we are more resilient at insisting that patients are either seen by us or seen in the Covid hub before deciding whether they need antibiotics, but I still have a hunch that we still are probably prescribing more readily than we previously were. GP14 Dec '20

I think that we were definitely using [antibiotics] a lot more indiscriminately because we just didn't know what we were dealing with and everyone was throwing the kitchen sink at it. GP10 Apr '21

It's probably back to where it was before. So I think when there was all the uncertainty earlier on in the pandemic we might have been prescribing antibiotics at a lower threshold for people, because we weren't reviewing them face to face. And if they were, and we're concerned about what, I know there's, there isn't any evidence for the antibiotics in Covid but of course we wouldn't know whether it was Covid or not, and if you had somebody who was high fever, short of breath and a cough, and you're trying to avoid them going into hospital, we'd give them antibiotics. Now that's changed and we either review them or we just do, where we just act as normal and give them self-care advice and ask them to get back to us if things are not getting better. GP08 May '21

…in the early pandemic, there was a bias towards prescribing. I think things settled, definitely things settled down over the winter. (…) I think, just because we started to go back to seeing, or having contact, with more patients with respiratory illness, the subtlety of distinguishing between whether somebody met the prescribing threshold or not, I think people started to move back to following their instincts a bit more, as they would have done previously. GP12 May '21

I think probably our threshold for prescribing for what is a viral infection was probably a lot lower than it would normally be, and, so I think probably at the beginning, I think that's probably back towards normal now. GP18 June '21

*Ambiguity about impact on antibiotic prescribing rates*

I'm sure that antibiotic prescribing for, say, cough or chest infection may have gone up. I haven't actually looked at it, but my perception would be that because we definitely did give some it… GP07 Nov '20

We've not seen [the data] since COVID, no. (…) I think our prescribing rates will have increased, particularly in spring, summer of this year. But I suspect they'll be coming back to almost back to a normal practice again. So, I suspect, well I hope, that they've fallen back the way they were before. GP08 Nov '20

I've just been looking at it, I mean the rate has gone down. I need to do a bit more (*inaudible*) but the early dataset that I've looked at, which is over a couple of months, so it's a couple of months around about a year ago, it's probably about 80% of what it was. I couldn't say, I couldn't give you that as a robust piece of information, just a general trend that's down. But I think the general trend of illness is down and the general trend of consultation numbers are down so it's difficult to know if that's practice behaviour or patient. There probably are a lot less respiratory illnesses this year than there have been in previous years as a result of, well there had to be really because transmission between people will be down. GP11 Nov '20

We do have a prescribing service that audit our antibiotics. I have to say, I don't know what happened to them, I haven't seen the report. (…) I suspect that, eyeballing it, overall, our antibiotic prescribing has gone down, simply because we haven't had many people this winter presenting with coughs and colds. I've got a chest infection doc, because there hasn't been any coughs or colds around. And then if they have Covid infection, we are not giving them antibiotics straight away. So, I would say that the overall number I suspect, when it does come out, will be lower than before. GP10 Apr '21

At the beginning, I would probably say we prescribed a lot less because we had so few contacts. People just weren't contacting us for a start. I'd say once contact increased, then we probably prescribed more because we didn't want to bring people in. Now most people are coming in, it's probably reduced again. GP15 May '21

*Antibiotic prescribing seen as less affected for other infections than RTIs*

I don't think COVID's changed a great deal of that [antibiotic prescribing for UTIs or skin infections], I think it's mainly for the respiratory tract infections that it changed. GP08 May '21

*Urinary tract infections (UTIs):*

For any other problems that are not respiratory related, I don't think the prescribing has changed that much. For example for UTIs they still have to drop a sample in, and if it's not hugely indicative of an infection we would still send the sample off before treatment. Or, if it's a simple cystitis, they will get a three-day supply of nitrofurantoin, and then only depending what the sensitivity would come back as, will they get then either an extension or a different course of antibiotics. GP04 Oct '20

…for UTIs it's more of a, that's something that's not necessarily going to change with the pandemic because there's nothing really that a face to face consultation is going to add to that anyway, so I think that's likely to be very similar to how it was pre pandemic. GP09 Nov '20

…for instance, suspected urine infections. In our local guidance there is quite clear criteria of when you should give antibiotics and when not to without needing to do a urine dip and things like that, and I think that's something that several of the GPs were used to just doing it based on, you know, symptoms alone and then there have been other groups of GPs and health professionals who have slightly more struggled to make the switch to not having the urine dip result as the decision maker on whether to prescribe or not. And things like that. I think probably there's been, I think personally, do I think? I think generally for things like urine infections, ear infections, my prescribing rates are probably about the same as they were before. GP12 Dec '20

Even pre-Covid, we were moving over to managing UTIs on the phone anyway. (…) we've got a practice protocol whereby if they have certain symptoms, so dysuria, nocturia and blood in the urine, and they are over 55, we prescribe a three days' course of antibiotics straight away. I think those are the criteria. If they have rigors, temperature or loin pain, then we probably have a lower threshold for seeing them face-to-face to make sure that they're not septic. And so, if we're not sure that those answers are not, questions are not no or yes, then we get them to drop off a urine sample, urinalysis and maybe *NCNS* depending on what the urinalysis says. So yeah, so I think that protocol has been around for a while, a lot of our, my colleagues were doing it in other surgeries in [area], pre-Covid, then we were just moving over to that when Covid struck. So, and there was some doctors in my surgery who weren't happy about that, they felt that they should be seeing everyone face-to-face pre-Covid. But then of course, the Covid hit, and we just thought, well this is the way we're doing it now. And I think most people are quite happy with it actually, most patients, because instead of coming in and waiting to be seen and having urine dipsticks, either we call them to say just drop a sample off and we'll call you when we've tested it. Or, just have three days' worth of antibiotics and if you're not better, call back. So, I'm not sure whether that's particularly Covid that's affected our antibiotic stewardship there particularly. GP10 Apr '21

*Skin infections:*

I don't think that it impacted on their care, for example, skin lesions. Or not skin, not lesions but skin conditions, skin lesions, we're always seeing skin lesions that were, that looked concerning for example, a melanoma, an early squamous cell carcinoma, we were always seeing these patients, for example, skin rashes, a lot more patients were sending in dermatological conditions. And where they might normally have come into see us with them, they were sending in pictures and for a lot of people that, we could manage that, actually, we could manage that well. GP08 Nov '20

Skin infections, I'd have to say I think there's, the using the text messaging services to receive photographs, and using the e-consult services to receive photographs, I think that's something that I think, again, people have got more confident at, over the course of the pandemic.  I think people have got better about saying to the patient, I want you to take a good photograph, or to take the two or three images that we need, so that we can properly assess this red patch on your leg, that may or may not be a skin infection, and things like that.  And I think again, that's probably, again that's changed in that again, early in the pandemic, people might have just prescribed anyway. Whereas actually now, people are much more confident about making sure they get good images through those electronic means, and then making an appropriate judgement.  Or, if there's lack of clarity, again more likely to bring patients in for a face to face now, than perhaps in the early pandemic. GP12 May '21

### Changing ways of working and consulting

#### *Move to remote consultations as leading to increased antibiotic prescribing*

…from March [2020], because sometimes the assessment is quite difficult on the phone or video.  So you err on the side of caution, because although you don't, antibiotics is not going to help with the COVID, but we couldn't listen to the patient's chest, we couldn't examine them, there were restrictions with the PPE, etc.  So we had to give them the antibiotics just to be on the safe side, to cover them for secondary antibacterial infections. GP03 Oct '20

We've had to adapt our way of working because we're having to work in a different way now doing a lot of consulting online or via telephone and so on.  In some ways it increases the stress levels because when you have to make a diagnosis or an interpretation of a patient's symptoms remotely, it introduces levels of uncertainty, I think, whereas perhaps we're not used to to that sort of extent... GP09 Nov '20

I do feel that we probably have prescribed more than we normally would, especially at the beginning, in March, April, May, [due to] the lack of face-to-face consultations, or moving everything online. GP10 Nov '20

With remote consultations I wouldn't have, I wouldn't have been comfortable to prescribe antibiotics a year ago.  Not often.  I mean it would happen occasionally if you knew a patient very well and you knew their history and their triggers and the natural progression of their illnesses, but if you didn't know all of that, then you'd want to get people in to examine them.  Now I'm more comfortable to, and I suspect that's because of a professional shift to, that maybe doctors now feel that a chest examination isn't definitely a must, whereas I suspect a year ago if you'd asked 100 doctors maybe the vast majority would say that a chest examination is a must when you are assessing someone respiratory.  Clearly it's desirable but now probably people will start saying well actually if you've got a very good history and a good understanding of it, perhaps there are situations when you could.  So I'm probably prescribing more antibiotics remotely than I would have in the past but equally I'm doing about 20 times more, sorry, no up to ten times more phone calls now than I was. GP11 Nov '20

I think there is a concern amongst several of the GPs including myself that there has been a slight increase in antibiotic prescribing as a result of the pandemic because of having that separation that the remote consultations create.  I think there is a concern that perhaps people are more likely to issue an antibiotic prescription than they might do if they had actually done an examination of the patient and can actually see them face to face. GP12 Nov '20

I think certainly in the early months of the pandemic antibiotic prescribing was higher than before where we were unsure of the risks of bringing patients into the surgery versus the risks of antimicrobial resistance or the risk of an untreated infection where a lot of people, a lot of prescribers were issuing antibiotics where previously we would have been more conservative and examined the patient first. GP14 Dec '20

*Gradually increase face-to-face consultations, return to more usual assessment & prescribing*

I think we're back to what we would, are back to our normal practice. And as I said, we're seeing more patients. So, we will assess patients, we're more likely to assess patients and we've got, and people that we, there's a better understanding of COVID, we, we're, I think people have got a better feel for the illness and I feel like, not that they're less frightened, but we didn't really know how soon the complications, *further complications* would develop or the risks of secondary bacterial pneumonias would be. And so, we're, I think people were seeing how it behaves a bit better, and so we're a bit more confident in assessing people now and knowing when to think it's, when it's likely to be bacterial versus a simple bio infection as we do normally. GP08 Nov '20

Now I think, now that there are more Covid hubs, I think as a group we are more resilient at insisting that patients are either seen by us or seen in the Covid hub before deciding whether they need antibiotics, but I still have a hunch that we still are probably prescribing more readily than we previously were. GP14 Dec '20

[Antibiotic prescribing is] probably back to where it was before. So I think in the, when there was all the uncertainty earlier on in the pandemic we might have been prescribing antibiotics at a lower threshold for people, because we weren't reviewing them face to face. (…) Now that's changed and we either review them or we just do, where we just act as normal and give them self-care advice and ask them to get back to us if things are not getting better. For example, if they're a younger person with a cough and a fever and they weren't, and they were eating and drinking and they weren't short of breath and they were stable. So I think we're back to our normal prescribing patterns and I think we're, I imagine we're prescribing, our rate of syndrome to antibiotic prescribing, or antibiotic to respiratory syndrome prescribing has dropped. GP08 May '21

We're doing it based on clinical needs rather than infection risks, to be honest. And now a lot of our, obviously all our, the vast majority of our vulnerable patients have had the vaccine and the vast majority of our patients over 40 have had the vaccine, so the risks even coming in, and all our staff has had the vaccine, so the risks of it are much lower. So basically we're doing it based on clinical need, and those that are not coming in actually we're dealing with just as efficiently on the phone with probably no, a face to face review for these patients would add very little anyway. GP08 May '21

as the pandemic has moved on, we've gone a little bit more back to normal, I suppose, being more strict, I think, in that we would expect patients to come in if there is a need for an antibiotic for tonsillitis, let's say, whereas at the very beginning of the pandemic we were perhaps a little more liberal to prevent patients coming, to come in, we were prescribing them with antibiotics over the phone. GP14 May '21

#### *Using Covid-19 clinics / hot hubs*

We've got a branch surgery where we saw patients who we didn't think had COVID, but may have had a fever, so potentially might have been exposed. Or if COVID was on the differential diagnosis, that further down, we had used the branch surgery as a hot surgery. And where we saw lots of people that might have been in contact of someone with COVID, but needed to be seen. GP08 Nov '20

We initially set up a red room within the practice which could be accessed separately from the other rooms and called that our CALM clinic and then, as cases seemed to be increasing we, and, I guess, our thresholds reduced about what we were concerned about, so pretty much anyone with a fever, we joined with our neighbouring practice. We're in a network with our neighbouring practice to run a CALM clinic in the middle of the building which was completely separate and wrote joint protocols to work that, so we had a very clear separate service seeing CALM patients run by a GP from their side for a session and a GP from our side for a session so we had full coverage across the day so that we could see our CALM Covid patients there and then we could still run a normal service in the rest of our parts of the building and have very clear infection control guidance with us and both infection control leads and the CCG support to run that. GP13 Dec '20

I am quite a tight prescriber on antibiotics for respiratory tract infections, so I am low risk when it comes to Covid infection complications, so I'll quite happily see our patients in our Covid tent to assess them if they couldn't go to the CALM Hub and I would assess before prescribing unless I knew the patient very well in which case I would treat them just as I would have done in 2019, I would treat them based on their history. So from my personal practice, isn't really any different I don't think to 2019, but perhaps that's because we're seeing, I'm seeing a lot less of these types of patients, they're selecting out, they're either getting their own swabs or they're already going to the CALM hubs through 119. GP14 Dec '20

Also lots of the coughs, colds, fevers were being seen either in the Covid hubs or having Covid swabs and just staying at home. GP14 Dec '20

### Changing patient presentations and workload

#### *Initially little patient contact, followed by increasing demand and workload*

In March, April probably May, it was just incredibly quiet and everything stopped basically. (…) basically, the routine part of general practice just, it wasn't, it just stopped for a couple of months, and then things changed after the lockdown ended the first time. GP08 Nov '20

Initially in April there was quite a lull in the number of patients that were calling us actually. It seemed that people were trying to avoid using the NHS at that point, but things have gradually got busier since then really. GP09 Nov '20

I think patient demand dropped as well, I think that was well documented, people just didn't contact us. And a lot of people were under the impression that we were closed for some reason, but we're not. And I think since the summer the workload has gradually increased, and I would say that in the last two months or so it's really back to normal, the demand is back to normal. (…) at the beginning, during the first wave, patients were, I think they were worried about coming in. So there were people who probably didn't call, because they didn't want to come into the surgery. I think they probably either thought that we were closed, I don't know why that is… GP10 Nov '20

March and April was very quiet. I think you'll hear that from other surgeries. I think the whole public wants to protect the NHS so we, I mean I wouldn't say it was completely quiet but it was very much, the volumes were very much lower than they are for instance now so we had that period in March and April. Then volumes gradually picked up. GP11 Nov '20

The demand from patients initially dropped a lot. After about four weeks we got a flurry of, sort of, increased demand, patients wanting to have things like their routine monitoring that had been delayed to take part. But then it was at the end of the first lockdown period that we then saw a very large exponential growth in patient demand. A lot of sort of unmet need where patients had basically put off contacting us about problems that had been ongoing even though we'd been very clear in advertising that, you know, we were available, you could phone, there were appointments available. But that, and that kind of high demand has continued basically since the lifting of lockdown in the summer. GP12 Dec '20

…there's all this unmet need, this massive unmet need at the moment, and secondary care, it's not functioning. And so we're getting a bit in the middle of that as a double whammy of having unmet need. So the, managing the unmet needs and then also having the unmet needs from the hospital because a lot of these clinics aren't functioning yet. GP08 May '21

I think we're much more ready to prescribe antibiotics because it's one way so you can not have to see the patient to make that decision. GP13 May '21

I think through the middle of the pandemic, after the first lockdown into the summer and autumn, it got back to sort of normal levels. And I think we've seen over the last few months that demand has gone through the roof and we're busier now than even at previous, pre Covid, winter levels of demand. It's even higher than that. GP18 June '21

*Fewer patients with seasonal infections (leading to fewer antibiotic prescriptions)*

The interesting thing is all the children, all the hot children disappeared, they still haven't reappeared… The parents just didn't call us about sick children. And I don't know, I think there's twofold, I think because of all the social distancing and all the schools stopping, actually stopped, reduced the incidence of viral infection in children, because they were not mixing with each other, so the *true* incidents probably went down. And secondly, I think parents are just, either don't want to bring their children into practice because they're worried about COVID, or they don't want to "bother" us, so the self-care became a lot better. So, I've literally seen, the number of children I have seen since March, I can count on the fingers of two hands, which is, from the list of 7,000, is amazing. GP10 Nov '20

It was very strange that we weren't seeing very many children so there was clearly a lot of more home management by parents which meant we weren't seeing so many pyrexial children and I don't really understand why, but I guess the benefit is that maybe patients were trying to manage their symptoms as best they can at home and often we know that children get better. GP13 Dec '20

I think, not due to our good stewardship, I think it's due to the fact that you, just no one came with coughs and colds that we've been prescribing very little. GP10 Apr '21

…because at the beginning there was, I think the number of patients in total calling in and coming in was lower, so I think we had a bit more time at that time as well, to maybe discuss those sort of things in more detail. GP18 June '21

*Using antibiotics for Covid-19 (changing evidence)*

I'm sure patients with COVID have been given antibiotics because we just weren't sure early on with what would be beneficial and what wouldn't. GP07 Nov ‘20

I'm sure that antibiotic prescribing for, say, cough or chest infection may have gone up. I haven't actually looked at it, but my perception would be that because we definitely did give some it, when COVID was poorly known and they were suggesting we do that. GP07 Nov ‘20

…since the pandemic, we'd probably be slightly erring on the side of caution and be saying, even though this is all the evidence given that we're not sure if you have COVID and we're not sure the, if actually then a secondary bacterial pneumonia or any pneumonia. So, yes, it may be slightly mixed messages because you'll say, well, we think it's a virus, but we're going to give you an antibiotic. So, it might slightly undermine what we've been trying to all say as a kind of party line, not party line, all be saying the same things up to that. So, we would probably give it more than we would have before COVID, which is probably obvious. GP07 Nov ‘20

I know there’s, there isn’t any evidence for the antibiotics in Covid but of course we wouldn’t know whether it was Covid or not, and if you had somebody who was high fever, short of breath and a cough, and you’re trying to avoid them going into hospital, we’d give them antibiotics. Now that’s changed and we either review them or we just do, where we just act as normal and give them self-care advice and ask them to get back to us if things are not getting better. GP08 May ‘21

…since November we, we’ve had the, I can’t remember when the Principle Study for antibiotics came out. I’m aware that we are prescribing far fewer antibiotics for chest infections over the phone, especially for possibly Covid patients. So, as I said, during the first wave when we didn’t know anything about it, the, I have to say personally, I don’t think I did it, but I observed other, some partners were like giving everybody antibiotics if they call up and they maybe Covid positive. So, I think that by and large has stopped, and then, are you aware that the reporting of the Principle Study (…) So they were looking at doxycycline and clarithromycin I think for suspected or possibly Covid patients, to see whether that reduced any symptoms or hospital admission rate, and that didn’t do anything. So, it, it’s not going to make any difference if you give them antibiotics early, which is logical really. So, I think, guided by that, we are prescribing far fewer antibiotics when people called and said that they’re Covid positive. GP10 Apr ‘21

During the first wave, when we didn’t know anything about Covid, we thought that, we did, tried in inverted commas, antibiotics to see whether it made any difference. That’s probably the only change I would describe Covid has affected me and my prescribing. But now that we know that antibiotics doesn’t make any difference to Covid, then I don’t think I’m going to change the way I work really. GP10 Apr ‘21

*Ambiguity over impact of COVID-19 on patient behaviours and expectations*

I think that [patients] want [antibiotics] more, because of the risk that they develop contagious respiratory infection, that they may be infected is the same. So there’s been a higher demand for them. GP01 Oct ‘20

I think people do understand that this is, COVID is a viral infection, and I don't know whether that's improved general public understanding of the use of antibiotics in viral infection. I think they do, that this probably highlighted the fact that COVID is a viral infection and therefore antibiotics don’t work, I think that that's a quite obvious take away lesson for the general public. GP10 Nov ‘20

…people seem to get the fact that Covid is a viral infection now. I think that's one thing that Covid's probably done in educating the public that there is a big difference between viral infection and bacterial infection. So, they all get it, they're not ringing us up to say, oh I'm, I've swabbed positive for Covid and I need some antibiotics. So, they don't do that, unlike when they've got flu or a cold, they say I need antibiotics. So, I suppose that, in that sense, it's probably done the public health a favour. Yeah, so yeah, I think patients know quite well that, if they've got Covid, antibiotics are not going to save them, yeah. GP10 Apr '21

Not really noticed any difference between early and late presentation (…) we don't know what we don't know, so there's people in, that listen to advice and don't call, we don't hear about. And my sense is there aren't many late presentations of acute respiratory illness, so I don't think people are deteriorating at home because they've been told antibiotics don't affect Covid and aren't effective against viruses and self-management. So my impression is there's not many people who delayed respiratory infections because of that message. But I don't know how many people have appropriately not contacted the practice because they're doing self-care now. GP08 May '21

…patients are probably the same as they've always been, they just want the right advice. I don't think they're assuming they're going to get antibiotics. I think they just want to make sure they're OK. I think patients had that period of feeling very fearful of knowing what their symptoms meant, very fearful if it was Covid or it wasn't. So I think knowing that they've had a swab or they're vaccinated, I think gives them more confidence, so I don't think they're expecting to get the antibiotics each time. GP13 May '21

I have found that patients, if anything, are managing themselves a little more before coming to us. So they might phone on day four or having a cough, once they've had a Covid swab, the result is back, then they get in touch, whereas beforehand, people would call us on day one or day two, so yeah, I've seen patients call on the surgery slightly later in the illness, but apart from that… GP14 Apr '21

…patients are a lot happier we're giving [antibiotics] to them now on a far lower threshold. That's the one thing I've seen, that they're, previously once you say no, it's a viral, they tend to get very grumpy and it'd be a longer phone call, longer consultation sorry, whereas now it's, you say, all right, it could be this, it could be that, could be viral, we have a lower threshold to prescribe, they're happier. They prefer it that way. Long term, for the general population and the public, what with antibiotic resistance, everyone is going to suffer, but people don't think like that, they think about themselves first and foremost. GP17 June '21

### Changing engagement with AMS strategies

#### *Mostly no (or limited) AMS initiatives in practices*

…in all the plans that we've done, and reorganisations we've done for COVID, we didn't include antibiotics in any of these plans. GP08 Nov '20

[Has the pandemic influenced how you view or approach antibiotics and antibiotic prescribing as a practice?] It's difficult to say as a practice, I think. I don't think I've, we've not really discussed it as a team, to be honest, how we prescribe antibiotics. (…) I don't think there's been any change in any strategy regarding antibiotic prescribing, no. We've each done our own thing, I think, individually as clinicians and there's not been any coordinated approach to it. GP09 Nov '20

We've basically just, myself and my colleague, have been reminding all the clinicians that antibiotic stewardship still applies, that the guidelines are still there. GP12 Dec ‘20

We’ve always had weekly practice meetings since pre Covid and that would involve GPs and the allied health professionals, so the ANPs, ECPs, so we’ve continued to use those opportunities. But then at the start of the pandemic we were actually having a daily morning briefing with all staff, because things were changing so quickly in terms of all the Covid guidance, (…) so that was another opportunity where we were able to remind people of things like antimicrobial stewardship and any other issues. Things like when the Covid, sort of, severity guidance and prescribing guidance came out from the CCG, that was how that was disseminated to staff in the practice, and things like that. That’s now been scaled back quite a long time ago now to only twice weekly… GP12 Dec ‘20

Thinking back over the last six months, there was one occasion at a practice meeting, where our Clinical Lead reminded everybody that mild viral illness, self-limiting infection should not have antibiotics prescribed. But that’s, as far as I can think back, that’s the only thing in the last six months or so that we’ve had, at a practice level. GP12 May ‘21

We sometimes have meetings, particularly the antibiotics that begin with C, so cipro, cefalexin, clindamycin and maybe discuss their prescribing (…) but we haven’t done anything like that for a year. We’ve hardly had any educational meetings for a year, partly because so many people are still working remotely, so it’s a bit difficult to actually organise. GP15 May ‘21

*CCG AMS work was mostly perceived as suspended during COVID-19*

…the Medicines Management team, for our local team, they've always got other things they want to talk to us about. I can't really remember antibiotics being one of their priorities recently. GP06 Nov ‘20

…every year we have a prescribing advisor who’s usually a clinical pharmacist employed by the CCG to look at our prescribing. And then we’re giving, usually given choices of three targets of various things, not always antibiotics, but sometimes it would be reducing macrolides, not prescribing, reducing the *floxacins* etc. I don’t know what’s happened to this year’s target, I can’t tell you. GP10 Apr ‘21

We’d normally sit down and look at [prescribing data] in more detail, and have an extensive chat with the CCG Prescribing Team about that. Obviously there’s more than, it also encompasses things like prescribing of strong pain killers, and various other drugs as well, and things like that. Because of the pandemic, we’ve not had that sit down and proper dig into the numbers. So I believe the document was circulated within the last 12 months, but that we just, it was for information only, rather than to actually discuss, and see if we needed to change practice in any way. GP12 May ‘21

I don’t think there are any specific things during the pandemic. I think it’s sort of, I think in our CCG there has always been quite a lot of support from them, yeah, from Medicines Management team about antibiotics and not prescribing particularly broad spectrum antibiotics… GP18 June ‘21

*Impact on, and use of, communication strategies*

…you communicate with patients differently when you can’t see them when you’re just talking over the phone so that’s changed because you haven’t got nonverbal cues, because you can’t see your patient, but I would explain things, no, in a similar way but I might say, if you’re not getting any better, *I'd* safety net, so encourage them to get back to us especially if I'm trying to monitor somebody that I think might have COVID, and then I would explain to them that I was giving them antibiotics because I wanted to make sure that they had, that I wasn't missing a secondary infection because I couldn't actually physically examine them or I've been discouraged from physically examining them. GP02 Oct '20

I do try to explain to patients why I do things or not do things, and why I prescribe antibiotics or not prescribe antibiotics, so I don't think COVID per se has influenced that, or the way we work. GP10 Nov '20

It's focussed the history component, I was just chatting with the other doctors and they'll all say the same, that a telephone or video approach does absolutely focus [on] the importance of understanding people's concerns, expectations, and that barrier between us and the patient and actually sort of, it can help in terms of having the discussion around what the normal expectations of an illness might be versus what would cause us worry and when we'd feel that people need to be seen. GP11 Nov '20

When people come in I think there's a feeling that they've made the effort to come in to surgery so they probably deserve, if they don't, if they're not given antibiotics perhaps you're fobbing them off. Whereas now because you're not, they're not making that effort to come in, they make an effort to book an appointment on the telephone lines, that's still a challenge for patients, but they're at home so you can generally have a much more mature conversation, a more expansive history, and get down to what they, to get down the, what their expectations and what their understandings of their illness is and understandings of what the normal symptoms they might expect for this type of illness are and how long they might expect it to go on. So it lends itself much better to have that conversation. GP11 Nov '20

…how we talk to patients about antibiotics, I think how we talk to them has probably still stayed the same, just the risk benefit conversation has changed, whereas, because there's a new risk in town basically. So I don't think it's really changed how we speak to them, it's just that, I suppose perhaps a little bit more specific perhaps with being clear that something is not this viral infection, before starting antibiotics perhaps. GP14 Dec '20

I think on the phone the safety netting is even more important, we can't see if they're actually taking the information in. But the advice I would give would still be the same and how I explain about antibiotics is still the same. GP16 May '21

It takes more time, you have to safety net more, you have to ask a lot more questions. GP18 June '21

*Impact on, and use of, delayed/back-up prescriptions*

I think we're probably doing more delayed prescriptions, so we're just advising [patients] if things get worse, get the prescription, but if you're better, then don't bother. So that is a quite useful strategy to have. GP01 Oct '20

…the threshold for antibiotic prescribing has gone down since the COVID. So we're not doing delayed prescriptions in COVID. GP03 Oct '20

I don't think I've even thought about [delayed prescriptions] since the pandemic, you've just got to prescribe straight away so yes, I guess it's removed that as an option completely. GP05 Nov '20

…the only difference has been that the threshold for saying to start the prescription might have been a bit lower. So, because, at the beginning, because of the uncertainty about the disease. So, we might have, we probably said, OK, we'll send a prescription round to the pharmacy for someone to collect, but normally we'd say, listen we could just carry on for a few days, we'd expect you to get better, whereas we might, now, like I said, if it's carrying on for a few days like this, start the antibiotics. GP08 Nov '20

…just to minimise footfall within the surgery we've gone to almost 100% electronic prescribing now, so that's just the way that we're having to do it now. (…) a prescription will be issued to the pharmacy and with clear instructions to the patient on the sort of circumstances in which they should go and collect it. So, I generally tell them, just I've done a prescription, it's there at the pharmacy but don't go and collect it unless you definitely need it. GP09 Nov '20

I suspect, thinking it through, we probably did a lot of delayed prescribing in March, April time, a lot more, sorry, because the resources just weren't really there, they weren't mature, they weren't set up. (…) I think we'd use it more as a, because you had so few options when you're speaking to people with respiratory symptoms at that time so one way of knowing whether it's a serious problem or not would have been to give antibiotics and see if they'd get better or not. GP11 Nov '20

I think it's harder for a patient to understand [an electronic DP] than if they were given the green prescription form and it just has a later date on it. GP12 Dec '20

…previously we may not have given any prescription, so our thresholds for giving a delayed prescription like our antibiotic prescribing has reduced in the sense of we're more likely to do it to try and reduce the chance of someone ringing back and saying the cough's got worse… GP13 Dec '20

Delayed antibiotic prescriptions have changed slightly. Not because of the pandemic but because we've now introduced electronic prescribing, and whereas previously we would perhaps give them a piece of paper and say, don't cash this in, now we have to work slightly differently in that we would say, for example, I'm going to put a prescription on your notes. If you need it, you need to get back in touch with us, so it's just another barrier. So I worry that some patients say, oh, can you just send it to the chemist? I'll collect it, I won't use it if I don't need it, so I think, so that has been a difference, but that's nothing really to do with the pandemic, that's to do with the change over to electronic prescribing… GP14 May '21

#### *Impact on, and use of, point-of-care CRP testing*

…when COVID came in and we weren't seeing anyone face to face, [point-of-care CRP testing] just got completely forgotten about. (…) I think in principle, it's a great tool to have as a clinician. The only difficulty is, as I say, if you've got a febrile illness or an illness suggesting infection, you're not going to get into the practice. So, we're not going to be testing you. GP06 Nov '20

…the COVID has taught us that, it really made us look at a lot of the conditions and why we're doing, why we're managing conditions in the way we're doing. So, and it's shown us that we can do things better and differently. So, I'm sure there is a way to do it [point-of-care CRP testing]. So how would you, to incorporate a CRP test. (…) So it needs, the patient needs to know to be seen at that specific time either by the clinician of some sort, (*inaudible*) a nurse, a healthcare assistant or a GP. So that's the bit in the process that they've got to think about. So, it needs to be timed. (…) So you've got to be able to book a patient, and it's not something they can just leave for, and then you can follow up afterwards, so you need to be able to book a patient in, and the patient needs a time to come and see somebody. And so then you've got to make that time available, rather than just, there is a difference in the processes, there's a difference between batch testing all your urine tests if you like, I could take that as a simple, most basic point of care test, versus this [point-of-care CRP test] which needs a process where the patient has got to see somebody, they'll have to know to turn up and get that done, and there's got to be a clinician available to do it there and then. And that's slightly different, not impossible, it's slightly different. (…) Yeah, and so, I guess that you're going to have to do it as part of the assessment, so it is part of the clinical assessment. (…) The other way to do it is to redesign the system for seeing respiratory patients and have them being triaged and being seen by our respiratory, the respiratory doctor for the day, if you like. And they would have, and then you could build the CRP into that system. So, they'd come in, they'd see somebody, they'd get a blood test done, they'd see a respiratory doctor of the day and (…) they'd have that CRP result when they saw them. That might be another way to do it. And that's not unfeasible because we are seeing respiratory patients a bit differently now, because [if] they've got COVID they've been seen in different places, they've been seen in the hot hubs, but also we're seeing, we're still seeing patients with cough and fevers at a different practice or at a branch practice that I told you about. The branch surgery that I mentioned earlier on. So, as I said, this is forcing us to do things differently, so it's not impossible, it' not impossible to include that in a different system. GP08 Nov '20

I don't know whether it would help in general, like a bit of a hot kid who's got viral infection, I'm not sure whether that will help. (…) if most of the consultations are mainly done remotely at the moment, then you can't do that anyway. But I'm thinking that, especially with all the vaccines on the horizon, at some point in the next couple of years we will go back to normal, in which case I think the CRP machine probably does have a big role to play. GP10 Nov '20

I think if you are choosing to do a CRP on someone, you're choosing to see them unless someone's going to go out to do their CRP at home and I think as an additional tool to someone who's got, who's unwell and got symptoms, it won't add much, in my opinion. I think those patients which we're thinking, gosh, they need some near patient testing, generally we're also wanting them to have a chest X-ray, possibly some blood gasses, and I don't think the CRP is definitive. I also think that as the result of the pandemic in our working, because our threshold ran to what excess we'd use, we're much more likely to say, well let's be on the side of caution and give you some antibiotics to cover you, if we think that they are safe to stay at home. GP13 Dec '20

Get more near patient testing perhaps in pharmacies which keep a lot of, we do deal with a lot of, I deal with a lot of sore throats and tonsillitis. Yeah, so I'm thinking maybe some near patient testing could help identify those patients that need, that would, who'd benefit from antibiotics and wouldn't. And I guess that maybe some simpler, some straightforward protocols which would include a near patient test could be done, that don't need GPs to do and interpret them. I think that would help make prescribing more consistent (…) the CRP testing adds, always adds time and that's the difficulty. So it's having a pathway where it's, it helps you make a decision but doesn't add to the consultation time. GP08 May '21

#### *Impact on, and use of, patient leaflets*

Obviously we can't really handing out leaflets anymore, so that's gone by the way. GP04 Oct '20

I think with COVID I probably have sent out more stuff because I'm not seeing [patients' face to face. So quite often I would say that there are sometimes when I do a consultation and then I will say, OK, I'll send you a leaflet, because now we're using a lot more texting as well. So, I can now text them a link to some sort of information or patient information leaflet, so I probably have, are using more in an electronic way, rather than the physical way, does that make sense? Rather than handing them out I'm sending them texts of information instead. GP10 Nov '20

Probably I use them more because what I'm doing now is texting a link to a patient rather than printing something out and giving it to them or posting it to them so because we use a texting system a lot more now, I probably am issuing more leaflets than previously. GP14 Dec '20

I think probably I used to print out the Treat Your Own Infection leaflets that exist for respiratory and urinary tract infection. And I don't know, practically, now I just find it easier to just send patients a text message with a link to the NHS website, rather than trying to send them a digital copy of those leaflets, and things like that. So I guess that's a change, in that I'd sort of, I'd just send them a link to the NHS website page now, rather than giving them a particular leaflet. GP12 May '21

*Impact on, and use of, guidelines*

We do have access to the local antibiotic prescribing guidelines, so I keep a copy on my desk top all the time so I can refer to the antibiotic guidelines very easily. And I've publicised that within the practice so that other clinicians do have access to that as well. So that's the main, my main source of help, I think, from an antibiotic point of view. GP09 Nov '20

There is obviously guidance for when it might be appropriate to prescribe antibiotics in a case of suspected COVID, and there is definitely, sort of, a much better correlation between the GPs deciding and making a clinical decision and recording a clinical decision whether somebody has mild, moderate or severe symptoms and then mirroring local CCG guidance on whether to prescribe antibiotics in those situations. GP12 Dec '20

I don't think the pandemic has changed our strategies as such, we still prescribe according to the [area] antibiotic network guidelines. GP14 Dec '20

*Impact on, and use of, clinical scores/templates*

If someone thinks they've got tonsillitis, I try to make an assessment of that, using either the Centor criteria or the FeverPAIN criteria to determine whether they may have a bacterial throat infection. Sometimes I've even asked them to send me a photograph of their throat, if they can do that. So that's sometimes a way of getting around the problem of doing a remote consultation. GP09 Nov '20

I certainly have used things like Centor or FeverPAIN (…) there have been occasional problems where the sort of fever element, because you're dealing with the remote consultation and people don't have a thermometer, it makes it harder to include that element in the scoring system. But for the most part most people actually are able to measure their temperature. So for the most part I'd generally be able to do things, either through a video consult, be able to do things like look at the back of someone's throat, or you know get someone to look in a mirror and then describe to you what's possible, so I would say most scoring systems I've been able to continue to use. GP12 Dec '20

I think the clinical score which has probably been used most has probably been used as part of our triage process and thinking about unwell patients with potential infections and potential sepsis and that seems to have worked quite well alongside assessing someone for Covid that we're using that score certainly a lot more. GP13 Dec '20

We use FeverPAIN score for tonsillitis, always have done for a long time, still do. Obviously, that is a little bit tricky to work out, can you see pus? Can you feel lymph nodes? So that's had to be, not fudged, but you have to be a little bit creative with that (…) previously, we'd always see patients, and look in their throats, a lot of the patients we're not physically seeing face to face, so we might do something over video or we might just ask a parent to see if we can get a perspective for what's going on. So we have to delegate that a little bit. GP14 May '20

…over the phone, but yeah I still use my normal ways, like to use FeverPAIN score, or UTI protocols (…) So with the FeverPAIN it's good usually to ask the patients all the questions and just put it into the algorithm. So I think it's how many days have you had symptoms, have you had cough, cold symptoms with it, have you got swollen glands, anything they don't know you can just put no answer. Fever in the last 24 hours. And then pus on tonsils, so that, and muscle aches, so that's all stuff you can actually just ask somebody over the phone, don't necessarily have to see them to be able to do that. And so yeah, that's quite useful because you can then also, either delayed prescribe or you can suggest to them when they need to phone back if symptoms haven't settled. GP16 May '21

### Shifting priorities

#### *Initial and current priorities related to the COVID-19 pandemic*

Our entire lives have been consumed with COVID planning and getting through the work. Because primary care is not an emergency service, so when COVID hit, we just went quiet. But I knew that what would happen was there would be a tsunami of pent up problems that will suddenly deluge us, and they did. And it's been painful, the amount of work we've had has been painful. And quite honestly, all those non COVID priorities have just slipped way down, because we're just dealing with workforce problems and capacity problems and staff going off sick. And it's just, our ability to stay on top of those other things, which ordinarily we'd want to tackle, like inappropriate prescribing or whatever, is just way down our list of priorities. We're just trying to get back on track now, but it's not going to be normal again for months. GP06 Nov '20

I think that it's so difficult to prioritize work. So, you know, we're discussing now, with the COVID vaccine rolling out, what are we going to not do (*laughs*) when we're doing the vaccines? So should we park this for a while, just forget about antibiotics? I'm not saying we do, I'm just saying, for instance, shall we stop doing that? They've already said maybe we should stop the medication review… GP10 Nov '20

At first we're, at the very beginning of the pandemic we were, we had, we'd accept, we were accepting much higher risk because we realised that at that point there was a lot of uncertainty about transmissibility and mortality, and the priority was to avoid transmission and to protect staff. GP08 May '21

I think we're all quite concerned about how busy it is now, in terms of just patient demand and how much worse that can get with the winter months. So I think that's going to be something where we're sort of concerned about. (…) trying to get the access because in terms of numbers of appointments we're the same as we were pre pandemic but the demand is just so much higher. GP18 June '21

#### *AMS not considered a priority during the pandemic*

Unfortunately, like a lot of things with COVID, it's put it, it's removed [AMS] from our top priority, unfortunately. And I think, again, I don't know if it's just our practice but other practices as well have maybe gone backwards a little bit as, becoming a little bit looser in our prescribing of antibiotics just because, as I said, we didn't have access to the patients and we were very keen to reduce a patient's need to come and see us (…) with COVID, as I say, [AMS] is not at the top of our priority list but hopefully, once we come out of COVID, then once again we can start prioritising what is a hugely important issue. GP02 Oct '20

It's been painful, the amount of work we've had has been painful. And quite honestly, all those non-COVID priorities have just slipped way down, because we're just dealing with workforce problems and capacity problems and staff going off sick. And it's just, our ability to stay on top of those other things, which ordinarily we'd want to tackle, like inappropriate prescribing or whatever, is just way down our list of priorities. GP06 Nov '20

What would make it more of a priority would be, well, I guess if there was, if we were, if there was, we had an adverse clinical outcome. So if someone wasn't prescribing properly or if we had cases where infections, we were not managing infections, we had significant events when infections weren't being managed properly. (…) that would probably be our stimulus for changing our, the way we, towards something to look at, to review our antibiotic prescribing. But I, we haven't had significant events where infections have been discussed. So I'm not saying it's never happened, I'm not saying it doesn't happen, I'm saying it doesn't seem to reach the threshold for being something to, there are adverse outcomes, an adverse outcome that I'm aware of. GP08 May '21

I just wanted to stay alive. (*laughs*) Working in the clinic, as I say, everything that was being done audit wise have just disappeared, completely disappeared. It'll come back, it will come back, but when you've got spikes in Covid rates locally, clinicians off sick, all your admin staff helping out with the vaccine clinics, it's all hands on deck at the moment. So, no type of audit, no matter how well intentioned, is going to get a look in. GP17 June '21

No, no-one's going to do it, at the moment, you'd be, anyone who tries to come in and starts saying, they're starting audits now, is just going to get completely shot down, or just get totally annihilated. Whoever it is that's going to just start saying that, I think no matter how well intentioned it is, anyone who comes in and starts saying, can you start doing these audits now? It will be, they will get complaints above their grade level just saying, what on earth are you playing at? We're trying to stay on top of this entire workload and you're coming out with this, that person isn't getting backed up by their own bosses, that's, it's a very difficult and challenging situation at the moment. Very difficult and challenging. GP17 June '21

*Difficulty planning due to uncertainty*

…it's a huge unknown, isn't it? And you don't really know from one day to the next one what's going to happen. GP04 Oct '20

The problem is also, we don't know what's going to happen with COVID for months. So, there's always a sense that we're still going to get battered in a few months' time, it's all going to get worse again. GP06 Nov '20

I don't think we've got any plans at the moment. No, it's so unpredictable isn't it? Because they're already talking about the fourth wave. And I think we'll just, I suppose it couldn't, I'd like to think it couldn't be any worse than the first wave. We were really flying blind this time last year, so I'd like to think that we wouldn't be as bad as that. GP10 Apr '21

*Lack of AMS-related plans but perceived value in re-engaging with it*

Everyone is still very much aware that [AMS is], well, it should be one the highest priorities to reduce inappropriate antibiotic prescribing. (…) I would hope once COVID is not the primary thing as it is for all of us at all times at the moment, then it should go back to continuing to be a forefront of what we're all trying to do. GP07 Nov '20

I think knowing that that has happened is important. So having very clear evidence that this has happened and people allowing themselves personal reflection about why and then hopefully having a personal challenge to alter what they do. I think overall, I think primary care does really well when tasked with changing practice. So if the CCG said to us, right, now, vaccinations have happened, care needs to go back to normal, we really need to sort out antibiotic prescribing, we'd like you to carry out a couple of audits, perhaps use a new tool and we'll put that as part of the prescribing incentive scheme for the following year so that your payment will be attached to that, I think that would probably work. I think GPs are very good at doing things that they have to do as part of being paid to do it and then sort of the powers within the practice which will alter people's management and carry out the audits will happen. I think if relying on individual clinicians appraising themselves without someone pointing out there's problems, probably won't make, it won't happen. So that's why I think it has to be at least a localised incentive scheme to partly reflect on antibiotic prescribing and secondly task us with reducing, well prescribing appropriately according to guidance and what the CCG would consider normal practice. GP13 Dec '20

Internally it's a project that we would like to do, is basically look at, do an internal audit on, well antimicrobial prescribing is one thing, but basically things like also frequency of, how frequently doctors and ANPs and ECPs refer patients to *statutory care*, and things like that, so that we've got slightly more rigorous data. To look at that, so that perhaps we can do some training, for people who, and support people who are higher rate prescribers, and things like that. But it's not something that we've had the time to do at the moment. GP12 May '21

I think we will get back to, when the annual CCG data comes out, the practice looking at that in more detail. But I think the sort of more formal audits of it, with exact numbers, is likely to take a lower priority, just purely because there is only so much time we've got for our administrative staff to do stuff. And I think that they know their time is going to be mostly taken up with the sort of changes in terms of managing the appointment book, and things like that, that are going to be happening, as we gradually come out of the pandemic. GP12 May '21

One of things that's useful that CCG have done previously that hasn't happened in this year so far and usually would have is that Oxfordshire CCG would usually pay for the GPs to go for an educational update. That hasn't happened this year. I don't know if that is going to be happening remotely or not, but there's been no talk of that and usually that would be attended by all the GPs, our paramedics, nurse prescriber and I presume that the physicians associates would go as well because they're consulting similar to we are, so that would be something as an opportunity for education… GP14 May '21

I think it would probably be more to do with getting the data back actually, just to see where we are and are there specific things we're doing differently to what we were doing before. GP18 June '21

*Some planning related to current priorities and potential changes to infection management*

The other way to do it is to redesign the system for seeing respiratory patients and have them being triaged and being seen by our respiratory, the respiratory doctor for the day, if you like. And they would have, and then you could build the CRP into that system. So, they'd come in, they'd see somebody, they'd get a blood test done, they'd see a respiratory doctor of the day and then they'd and then we would include, then that would be, they'd have that CRP result when they saw them. That might be another way to do it. And that's not unfeasible because we are seeing respiratory patients a bit differently now, because [if] they've got COVID they've been seen in different places, they've been seen in the hot hubs, but also we're seeing, we're still seeing patients with cough and fevers at a different practice or at a branch practice (…) So, as I said, this is forcing us to do things differently, so it's not impossible, it' not impossible to include that in a different system. (…) It's got to be something that's done on the routine without having to be thought about. It's just going to be part of the routine. As soon as somebody's got to stop and think about it, it won't get done. GP08 Nov '21

…the amount of work we've had in the last three months has been so utterly overwhelming, that I've just got a whole long list of prescribing things. Not necessarily antibiotic related, but loads of things that need dealing with, which just the chance of getting through them is, at the moment is minimal. We're supposed to be getting some QCN pharmacists starting shortly. GP06 Nov '20

We're changing, we're trying to change the way GPs work just so that they can spend more time seeing patients and less time doing the tsunami of admin that we were getting just now from all quarters. GP08 May '21

We've also got more permanent staff, which has again, helps improve patient continuity, rather than having lots of locums, and things like that. So I think that's the big thing, is sort of continuity of care for patients with a specific doctor has improved, and things like that. We're still operating on a telephone triage model first, and then bringing in patients for face to face where that's necessary and appropriate. (…) We're currently liaising with our Patient Participation Group over sort of what the future of our appointment book will look like. (…) from the positives that have come out of this pandemic in terms of the use of these new technologies, and people gaining confidence in terms of doing all the remote consultations. So that we don't take a step backwards from where that has been advantageous, and maybe preferable, for some of our patients, so that's something that's going on. We are trying to look also at the relative workload of kind of minor illness, urgent care presentations, versus more chronic disease presentations. (…) So that's in terms of looking at what balance do we need between GP workforce and then ANPs, ECPs, first contact physiotherapists, mental health workers, things like that. So we're trying to gather some data about that, so, in terms of workforce planning. GP12 May '21

I think our main plan is to try and get the flu programme sorted as early as possible, so we'll try, because obviously we don't know what's going to happen with booster immunisations for Covid and how that's going to work. GP13 May '21

I think our supportive staff like the pharmacists have a big role potentially to help us with not only prescribing, but also potentially working across the two practices and having consistency in the way we do things. I think the primary care network and the staff that they are hoping to employ will make a difference with trying to manage our infected type symptoms so, supposedly we're hoping to recruit a clinician's associate, a paramedic, which I think would also help. I think for the practice we recognise that minor is perhaps, it's a misterm but the type of work that team does and maybe acute care team would be a better title, so we probably will look at trying to utilise that team better.

I mean I hope that we will be able to see more patients face to face, but then we have to speak to less people to be able to do that. And I guess the only way that can happen is either by employing more staff, or thinking of other ways to deal with the demand. So I think part of that will be with the training to redirect patients to other *sources* of health, it doesn't necessarily need to be a GP or a nurse or one of our staff, it could be a pharmacist in the community, it could be online, it could be some other way to actually get them the help and information, so to decrease the number of patients we have to call. I, yeah, no, my hope is that we can see more and more patients face to face, and, but without those changes I can't see how it's possible to be able to achieve because there's not enough time in the day. GP13 May '21

I guess my feeling is that a lot of antibiotics could potentially be done in the community by pharmacists, certainly Trimethoprim for UTIs, I don't think really needs a GP to consider, and I think the joined up care of our pharmacy team, colleagues, potentially there are some things that they could do. GP13 May '21

So we've got that workload in hand, so our healthcare assistants who do the baseline assessments for the long term condition reviews, the bloods, the blood pressure, the weight, asking about alcohol, asking about smoking, they are working flat out now to get that workload up to where it should be and try and get ahead of where we would usually be in September, October time. GP14 May '21

So one of the things that I think we could do is now that we're using physicians associates and non-doctor, or non-prescribing people, to assess for minor illness, which is quite a high prescribing area, I think that we should have more templates that our surgery uses. So for example, I know there is a template for a FeverPAIN score, it's rarely used. We don't have a template for putting the information around UTI into our records, then I think that would make prescribing and coding of infections much clearer. It's just not something that, so our physicians associates are very junior to the role and to primary care, so it's not something that they know how to do, (…) I think if we could have more minor illness templates, that would make our coding of the minor illness and our prudent prescribing tidier. GP14 May '21

In terms of staffing, we've taken on extra staff so we're getting, the whole *site* is growing, so we were due to take on at least one other GP anyway but we're getting two GPs, we're getting an extra nurse, we've already taken on extra healthcare assistant because of meeting demand. GP14 May '21

I think it will be more like it is now actually. I think there will be a lot more remote and telephone stuff done so that's changed. The reason I think it's going to happen is because it was happening anyway and so it was the path of travel we were already on and it's been massively accelerated by the time of Covid and I don't think it's necessarily a bad thing. GP15 May '21
